# Rethinking the residual approach: Leveraging machine learning to operationalize cognitive resilience in Alzheimer’s disease

**DOI:** 10.1101/2024.08.19.24312256

**Authors:** Colin Birkenbihl, Madison Cuppels, Rory T. Boyle, Hannah M. Klinger, Oliver Langford, Gillian T. Coughlan, Michael J. Properzi, Jasmeer Chhatwal, Julie C. Price, Aaron P. Schultz, Dorene M. Rentz, Rebecca E. Amariglio, Keith A. Johnson, Rebecca F. Gottesman, Shubhabrata Mukherjee, Paul Maruff, Yen Ying Lim, Colin L. Masters, Alexa Beiser, Susan M. Resnick, Timothy M. Hughes, Samantha Burnham, Ilke Tunali, Susan Landau, Ann D. Cohen, Sterling C. Johnson, Tobey J. Betthauser, Sudha Seshadri, Samuel N. Lockhart, Sid E. O’Bryant, Prashanthi Vemuri, Reisa A. Sperling, Timothy J. Hohman, Michael C. Donohue, Rachel F. Buckley

## Abstract

Cognitive resilience describes the phenomenon of individuals evading cognitive decline despite prominent Alzheimer’s disease neuropathology. Operationalization and measurement of this latent construct is non-trivial as it cannot be directly observed. The residual approach has been widely applied to estimate CR, where the degree of resilience is estimated through a linear model’s residuals. We demonstrate that this approach makes specific, uncontrollable assumptions and likely leads to biased and erroneous resilience estimates. We propose an alternative strategy which overcomes the standard approach’s limitations using machine learning principles. Our proposed approach makes fewer assumptions about the data and construct to be measured and achieves better estimation accuracy on simulated ground-truth data.

## Introduction

Cognitive decline represents the cardinal clinical symptom of Alzheimer’s disease (AD), its manifestation across patients is heterogeneous^1,2^. Some patients exhibit rapid cognitive decline, while others do not show any clinical progression, despite showing similar levels of AD pathology^3^. This phenomenon is termed cognitive resilience (CR)^4^. CR is described as an individual exhibiting less cognitive decline than expected given their individual characteristics and pathological burden^5^. These characteristics can include social determinants of health, genetic risk, and AD pathology biomarker levels. The ‘expectation’ here references the cognitive performance if it would remain unaltered by CR. Constructs like CR cannot be observed or measured directly but manifest as a latent construct that alters an observable outcome (here, cognitive performance)^6^. Given that they potentially offer insights into protective mechanisms or intervention opportunities, however, the operationalization of latent constructs, such as CR, is paramount.

Previous studies of CR often circumvented the challenge of direct measurement by focusing investigation on the determinants of reduced cognitive decline instead of operationalizing resilience itself^7,8^. Alternatively, many studies considered a threshold on cognitive scores to define the point at which individuals are presumed to be resilient^9^. There are multiple disadvantages to the latter approach; for example, it reduces the latent construct of resilience to a binary condition even though it likely represents a continuous property that manifests over time.

A well-adopted alternative is the residual approach^10–14^. Here, a continuous measure of resilience is derived by calculating the deviation of an individual’s cognitive performance from the prediction of a linear regression model. The regression model is supposed to predict the expectation for an individual, such that positive residuals denote CR as the observed outcome is better than their prediction. Concerns about this standard residual approach have been published^15^ and the necessity for a better understanding of the residual approach was argued in a recent comprehensive review about the results achieved with it^16^.

In this work, we address the measurement of latent constructs, such as CR, using residual approaches. We explore properties and fail states of the standard residual approach and based on these limitations propose a novel inverse learning method that expands on the residual approach. The presented framework uses inverse supervised learning^17^ to predict the ‘expected’ outcomes of individuals and then corrects the prediction error that remains in the resulting CR estimates. Besides CR, it can operationalize any construct that can be described as a deviation from an expectation, for example, also pathological resistance. When compared to the standard residual approach, the inverse learning approach requires significantly fewer assumptions about the data to achieve accurate estimates. Using simulated data with a known ground-truth, we highlight the differences in the standard and proposed residual approaches and assess their limitations and estimation accuracy.

## Methods

### Problem definition

The aim of the residual approach is to measure by how much an individual’s cognitive function is improved by the latent construct of CR. The only entity that can be directly measured in this context is the observed cognition, *cog* _*exp*_, that is potentially altered by CR:

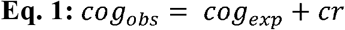

Here, *cog*_*exp*_ denotes the expectation, meaning the cognitive function an individual would have if they were unaffected by CR. *cr* represents the shift in cognition caused by CR. The residual approach aims to estimate *cr* given some data matrix *X* and *cog*_*obs*_. Notably, the same principles apply to any latent construct that can be described as the deviation from an expectation.

### Simulating data with a known ground-truth

As individuals’ true values of CR remain latent in real data, there exists no ground-truth against which different methods’ estimation performances can be compared to assess their quality. Accordingly, for our experiments, we simulated data and a known ground-truth analogous to Eq. 1, by i) simulating the expected cognition of samples and ii) altering it with a known CR component. For the remainder of this work, we will still refer to these simulated data as (expected) cognition and CR, for simplicity.

We generated a data matrix *X, X* ∈ ℝ^1000×27^, where the rows of *X* contained the samples and columns contained variables. We chose to simulate 27 variables because this allowed us to have sufficiently many expectation-associated, CR-associated, and uninformative variables that we could alter in our experiments outlined below. Next, we split *X* into a set of samples that were not resilient (i.e., resembled the expected outcome), *X*_*exp*_, and another set which was resilient, *X*_*cr*_.

Analogous to Eq. 1, the observed cognition, *cog*_*exp*_ and *cog*_*cr*_, corresponding to the samples in *X*_*exp*_ and *X*_*cr*_, respectively, can be simulated as

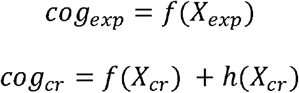

where *f* models the relationship between variables in *X* and *cog*_*exp*_ in Eq. 1 and *h* models the effect of variables in *X* on *cr. f* and *h* can be any linear or non-linear transformation. We parameterized *f* and *h* with two disjoint sets of nine variables of the 27 variables in *X*, respectively. A third set of nine variables remained unrelated to both transformations.

Unless stated otherwise, we used the following parameters to simulate data for our experiments: We simulated one third of the samples in *X* (i.e., 333 samples) to resemble the expectation (i.e., not adding a CR component to their outcome). This fraction was chosen to allow for meaningful simulations and the effect of altering it is explored in our experiments outlined below. Variables were generated to be uncorrelated. For simplicity, we assumed a linear relationship between *X* and both, *cog*_*exp*_ and *cr*, respectively. In this case, both *f* and *h* computed the dot product of their respective nine informative variables in *X* with a vector of coefficients, where all informative variables were assigned the same non-zero coefficient.

### Simulation experiments

We compared the standard residual approach to our proposed inverse learning approach across different experimental settings, where we independently altered the a) available predictors, b) correlation between predictors, c) prevalence of CR in the samples, d) and the relationship between predictors, cognition, and CR.

We evaluated how the selection of predictor variables affected the standard approach and our proposed approach. Successively, we include one third of the variables that were either informative for the expectation (*cog*_*exp*_ in Eq. 1) or CR (*cr* in Eq. 1) as predictors to the model. We then applied both residual approaches using all combinations of removed predictors.

In real cohort data, variables are often correlated, for example age and pathology, or education-level and cognition. We therefore evaluated how varying degrees of correlation between expectation-informative variables and CR-related variables affected the estimation of CR. To allow for precisely tuning the correlation between variables, we sampled the data matrix from a multivariate standard Gaussian distribution for this experiment. The corresponding covariance matrix was constructed such that simulated correlation ranged from uncorrelated to perfect collinearity.

In real AD data, the relationship between variables, cognition, and CR are probably non-linear^18^. We therefore assessed the estimation performance of the residual approach when modelling *f* and *h* as non-linear functions. To obtain a non-linear relationship between *X* and the expectation, we simulated data following Friedman et al. as implemented in scikit-learn ^19^. The explicit non-linear forms are presented in the Supplementary Material. A non-linear CR function *h* was simulated using an artificial neural network with random weights where the first layer was adjusted to ensure only select predictors were informative.

### Implementing the standard approach

We implemented the standard residual approach as in previous studies^10–14^: The linear regression model was fit on all the samples in *X*. The predictors changed depending on the experimental set-up described above. CR estimates were achieved by subtracting the prediction made by the linear model from the simulated observed value. We assessed the model fit using either the explained variance or coefficient of determination (R^2^).

### Training the predictive models of the inverse learning approach

In our inverse learning approach, any regression algorithm is viable for building the ‘expectation’ model that predicts the expectation component of the observed cognition (*cog*_*exp*_ in Eq.1). In our experiments, we used multivariable linear regression and extreme gradient boosting (xgboost), a non-linear approach based on an ensemble of decision trees. While any algorithm could be used here, we opted for xgboost because it is known to perform well on tabular data^20^. All models were trained to predict the observed cognition *cog* _*exp*_, of the samples unaffected by CR, *X*_*exp*_. To test the expectation model’s predictive performance, we conducted 5 times repeated 5-fold cross-validation. For details about hyperparameter tuning for the xgboost models see the Supplementary Material.

While any regression algorithm could be used for the ‘error correction’ model, in our experiments, we used a linear regression. The same predictors and samples as in the expectation model were used for model fitting. The training targets were the prediction errors of the expectation model for each sample gained during its cross-validation.

## Results

### The inverse learning approach

Our proposed inverse learning approach expands on the concept of the residual approach. Accordingly, CR is still operationalized as a residual of a sample’s observed cognition with respect to a predicted ‘expected’ cognition (from here only referred to as expectation). Our proposed approach works as follows: We first build a training dataset by selecting samples that follow a predefined definition of the expectation, deliberately aiming to exclude samples from the training data that could represent cognitive resilience. Based on this training dataset, we then build an expectation model that predicts the corresponding observed cognition which is, by definition, reflecting the expectation **(Figure 1A)**. This expectation model can now be applied to all remaining samples, which may or may not be cognitively resilient, to extract their residuals (**Figure 1B)**. Further, we introduce an ‘error correction’ model that aims to improve the accuracy of achieved CR estimates by removing the expectation model’s innate error from the calculated residuals **(Figure 1C)**.

**Figure 1:**
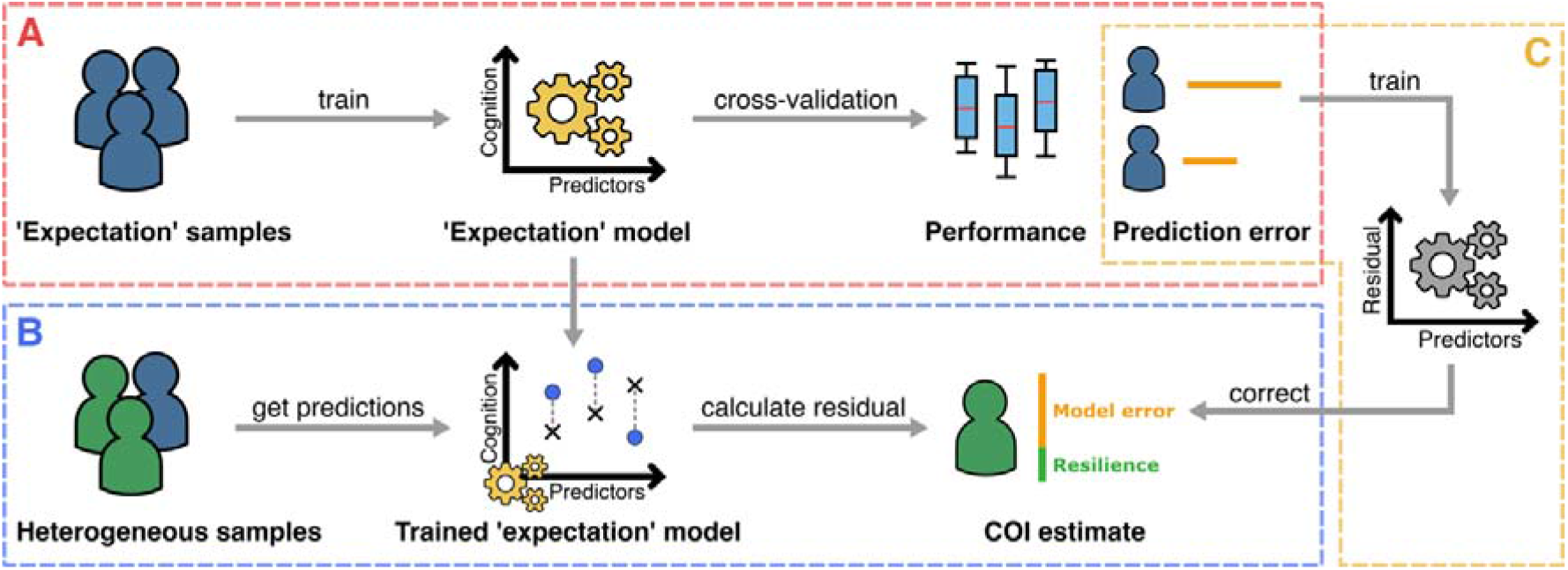
The proposed inverse learning approach for measuring cognitive resilience (CR) or other latent constructs that can be defined as a deviation from an ‘expectation’. **A**: Training a model to predict the expectation for a sample and evaluate the predictive accuracy. For this, only samples are used that follow a definition of what is ‘expected’. **B**: Applying the trained expectation model to samples that could be affected by the construct of interest to estimate it using the calculated residuals. **C**: An ‘error correction’ model that learns to remove the expectation models innate prediction error based on the errors in A and is subsequently applied to remove the error from the residuals obtained in B.

### Investigating the performance and fail states of the approaches

#### Effect of predictor selection

Variables used as predictors in residual approaches can be tied to both, CR, and the expectation component of a sample’s observed cognition (i.e., informative for both and in Eq. 3). We evaluated the effect of incorporating different predictors into our expectation model and the standard approach for estimating the sought-after simulated CR.

Including predictors informative for the expectation component of the observed outcome improved CR estimation accuracy of both approaches and better predicted the expectation (**Figure 2A & B**). Investigating the effect of including variables explaining the CR component of the observed cognition, we observed disparate results in each approach. For the standard approach, the achieved residuals became increasingly biased with the inclusion of more CR-informative predictors (**Figure 2C;** the bias is indicated by a systematic deviation from the diagonal line in the figure). Only when none of the included predictors were associated with CR, the standard approach’s residuals placed close to the simulated CR ground-truth (i.e., the optimal outcome). By contrast, the estimation accuracy remained stable for the inverse learning approach, irrespective of the number of CR associated predictors included (**Figure 2D**). In conclusion, the standard approach requires all predictors explaining the expectation component of the observed cognition but none of the CR associated variables to achieve the best CR estimates. The inverse learning approach, on the other hand, solely depends on the inclusion of variables tied to the expectation component, while being unaffected by the inclusion of other variables.

**Figure 2:**
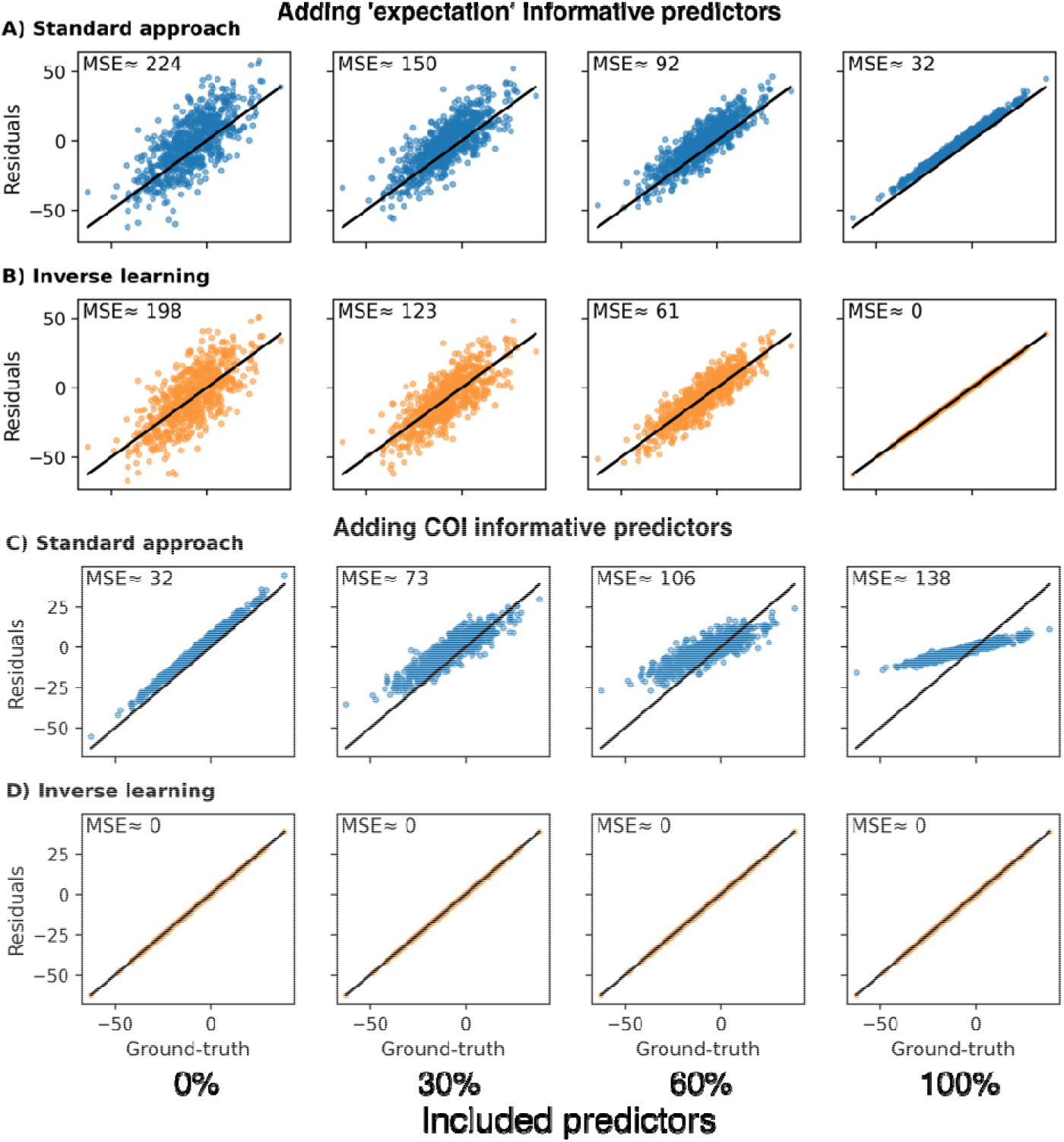
Estimation of CR using the standard and inverse learning approach while including varying proportions of informative predictors for either the CR or ‘expectation’ component of the observed cognition. **A, B:** Estimation without including any CR-informative predictors and varying proportions of ‘expectation’-informative predictors for the standard approach (A) and inverse learning approach (B). **C, D:** Estimation while including all ‘expectation’-informative predictors and varying proportions of CR-informative predictors for the standard approach (C) and inverse learning approach (D). MSE: Mean squared error between the simulated CR ground-truth and estimated CR.

#### Effect of correlation between ‘expectation’-informative and CR-informative predictors

To investigate the effect of variable correlation on the residual approaches, we simulated data with varying degrees of linear dependency across variables linked to both components. In this analysis, both the standard approach and inverse learning approach only included expectation predictors.

Despite not having access to CR associated variables, the standard approach still extracted biased residuals for increasing levels of variable correlation (**Figure 3A**, the bias is again indicated by a systematic shift of form the diagonal). In presence of correlation with CR-informative predictors, the inclusion of expectation associated predictors alone sufficed to reproduce patterns similar to those observed when directly including CR predictors in the standard approach (**Figure 2C**). The estimates for the inverse learning approach remained unchanged irrespective of the induced correlation (**Figure 3B**).

**Figure 3:**
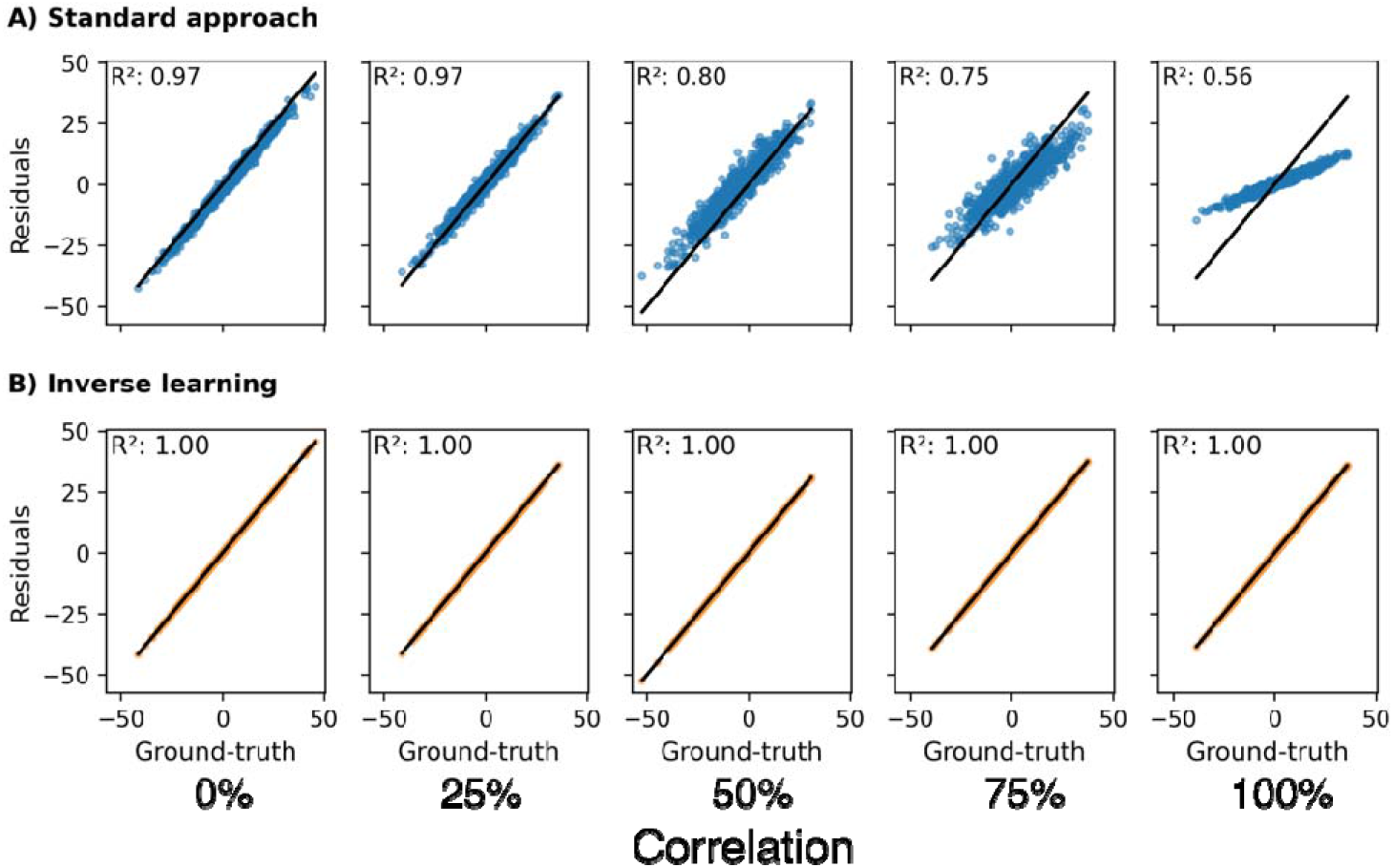
The effect of correlation between ‘expectation’-informative predictors and CR-informative predictors on the accuracy of CR estimates. 0% correlation means variables were entirely uncorrelated and 100% denotes perfect collinearity. All ‘expectation’-informative predictors were included in the models while CR predictors were omitted. Different from other plots in this article, the data values shown here differed across columns as simulating different correlations led to changing values. **A:** Estimation using the standard approach. **B:** Estimation using the inverse learning approach. **R**^**2**^: Coefficient of determination.

#### Effect of CR prevalence

In the standard approach, we found that higher prevalence of CR among the samples led to less accurate CR estimates (**Figure 4**). In the presence of variable correlation or when CR-informative predictors were included in the approach, higher prevalence led to vanishing residuals and, consequently, systematic underestimation of CR (**Figure 4A**). Even when no information about CR was accessible to the model, a systematic shift in the estimations occurred, biasing the achieved estimates (**Figure 4B**). The inverse learning approach, on the other hand, cannot be affected by the CR prevalence due to the exclusion of CR-affected samples from the expectation model training (assuming enough samples to train a reliable model remain).

**Figure 4:**
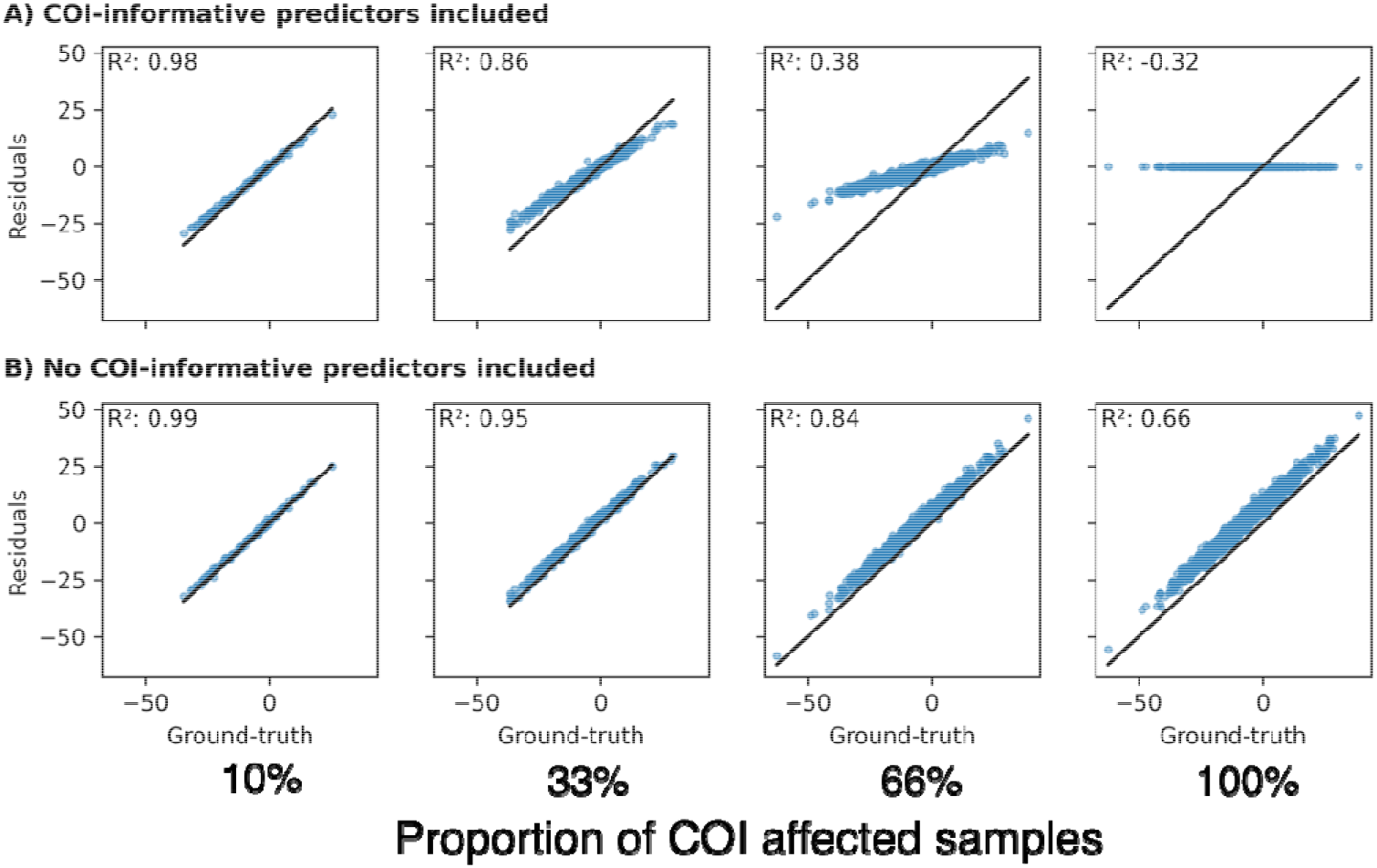
The effect of the CR prevalence in the dataset onto residual estimates achieved via the standard approach. **A**: All ‘expectation’-informative variables and CR-informative variables were included as predictors. **B**: Only ‘expectation’-informative predictors were included. R^2^: Coefficient of determination.

#### Effect of non-linear associations between ‘expectation’, CR, and predictors

Even with a non-linear relationship between predictors and CR, the standard approach produced biased CR estimates since the model still learned to explain some of CR and thus removed it from the residual (**Figure 5A**). As expected, in this scenario, a linear expectation model outperformed a non-linear one in the inverse learning approach, as the simulated expectation was linearly dependent on the predictors. When we tested the approaches on a non-linear expectation and a linear CR relationship, the standard approach failed to predict a reliable expectation and achieved poor CR estimates (**Figure 5B**). The non-linear inverse learning approach (**Figure 5B XGBoost**) performed best, after its estimates were corrected using the ‘error correction’ model.

**Figure 5:**
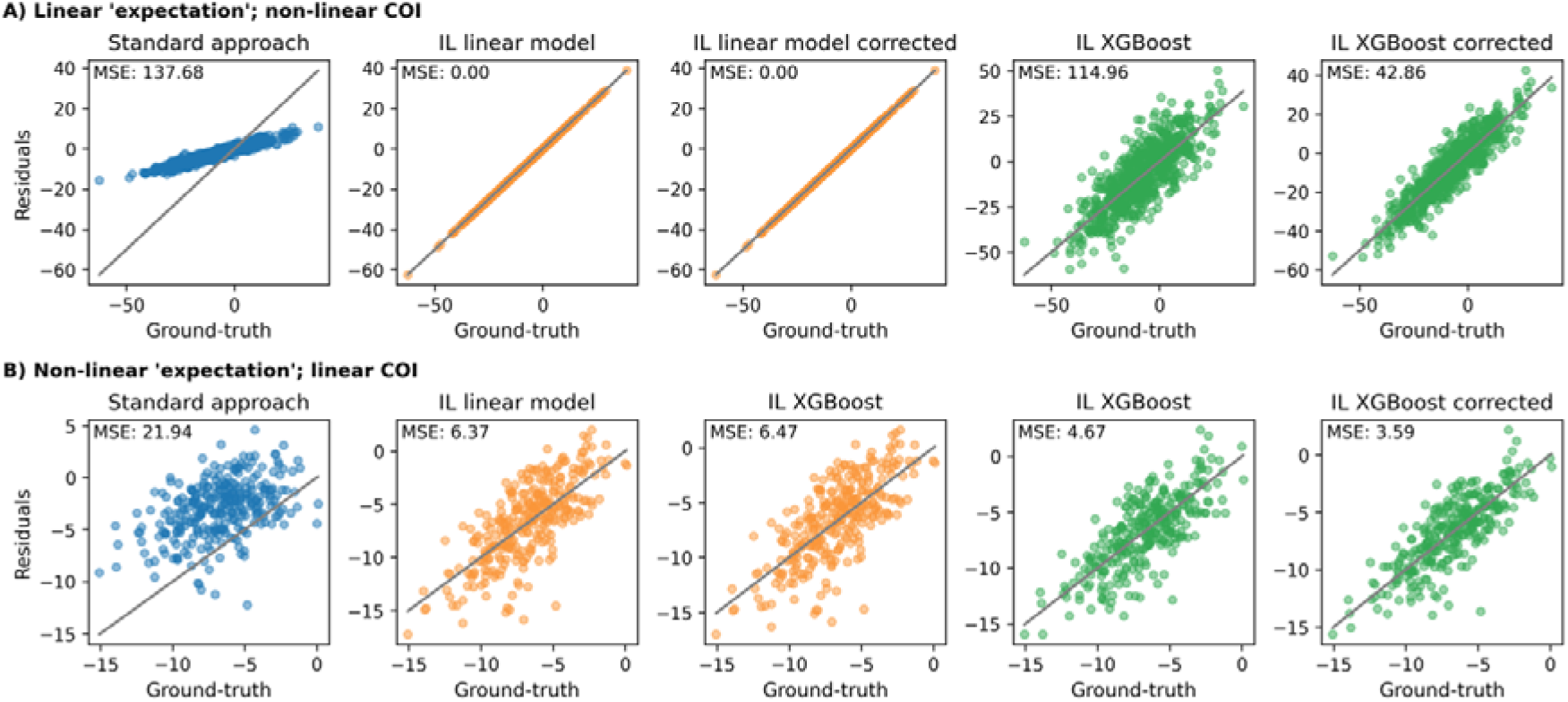
Comparing the residual approaches under different functional forms underlying the relationship between ‘expectation’ predictors, CR predictors and the observed outcome, respectively. Both approaches had access to all predictors. **A**: The ‘expectation’ component depended linearly on the predictors and CR component was simulated under a non-linear relationship. **B**: A non-linear relationship between predictors and the ‘expectation’ component was simulated and a linear relationship for CR component. Corrected: Estimates after running the ‘error correction’ model. IL: Inverse learning. MSE: Mean squared error.

In summary, when challenged across all the tested scenarios, the inverse learning approach achieved lower estimation error compared to the standard approach. The additional ‘error correction’ model substantially improved the predictive performance of non-linear expectation models (Figure 5) but did not aid linear expectation models.

## Discussion

Our proposed inverse learning strategy represents a framework for measuring CR and any latent construct that can be described as a deviation from an unobservable ‘expected’ outcome via a predictive model’s residual. It makes fewer assumptions that the standard residual approach about the data and relationships between predictors and CR. Consequently, this will lead to more accurate, individual estimates of CR in most scenarios. The conceptual differences between the standard approach and our inverse learning approach are graphically explained in Figure S1.

The main challenge in applying the standard residual approach appropriately is retaining CR information in the residual of a predictive model while simultaneously minimizing model error^21^. Our results show that, to achieve accurate estimates, the standard approach requires the following strong assumptions: 1) that the linear model accurately predicts the expectation, 2) that no CR-informative predictors are used, 3) that used predictors are uncorrelated with CR-informative variables, and 4) that the prevalence of CR among the investigated samples is low. Navigating these assumptions is challenging as they cannot be observed in real data. Often, there is no *a priori* knowledge about the relationship between variables, the observed outcome, and CR. It further remains unknown how many samples are affected by CR before estimating it. In contrast to the standard approach, the inverse learning approach only requires assembling a training dataset containing samples that resemble the expectation and an optimization of the expectation model adhering to machine learning best practices.

In both discussed residual approaches, the predicted expectation represents a crucial reference point to calculate the achieved residual. Predicting it inaccurately will add error to the residual and limit its reliability as a CR estimate. Therefore, it is vital that the models used in either approach achieve high predictive accuracy. Optimizing the performance of the linear regression in the standard approach, however, is non-trivial. To achieve the best predictions, meaningful predictors are required and all variables that can potentially improve the predictive accuracy should be incorporated. However, CR estimates made by the standard approach become severely biased if predictors are included that explain the CR component of the observed cognition. This is because the model learns to explain the effect on cognition caused by CR and, thus, CR information will be ‘learned out’ of the achieved residual. This poses a dilemma, as valuable predictors must be excluded from the model to retain CR information in the residual, at expense of predictive accuracy. One such example would be education which is relevant for both, the expected cognitive performance and cognitive resilience^8^. Consequently, the achieved residuals will be erroneous and inaccurate estimates of CR in most real-world applications. Our proposed inverse learning approach is not subject to this restriction. Due to building the expectation model only on samples reflecting the expectation, it is unable to learn associations between its predictors and CR. Consequently, all variables can be included into the model to maximize its predictive performance without fearing a loss of CR information in the residual. Additionally, more complex predictive models than a linear regression could be utilized to achieve the best predictive performance.

With the standard approach, the reliability of the predicted expectation cannot be directly assessed. Although the fit of the regression line can be measured (commonly done by looking at the explained variance of the observed outcome), in many real-world applications, the model likely incorporates information about CR through either correlation or inclusion of CR-informative predictors. Consequently, the model fit does not reflect how well the true expectation is predicted for an individual but how well it fitted the overall cognition. In our proposed inverse learning approach, a direct evaluation of the expectation model’s predictive performance is conducted via cross-validation. Additionally, an external validation can be performed to assess generalizability. While the linear models in the standard approach could be validated in theory, the resulting model performance would still not reflect its ability to predict the expectation of an individual but the overall observed cognition.

Despite the evident problems in the standard residual approach, many of its findings in CR research seem sensible from a neurological perspective^16^. While it is probable that many of these studies have been affected by some of the conceptual problems in the standard approach, it highlights the general robustness of the standard approach. Our experiments show that, even when it achieved very poor performance in predicting the expectation and included no informative predictors, the general trend of the residuals (higher residual equals greater resilience) remained true. This implies that the achieved residuals can provide some degree of insight related to associations between variables and CR, despite being inaccurate estimators of the actual resilience. Exact effect size estimates such as regression coefficients, however, are more uncertain.

When using real data of patients with neurodegenerative diseases, it is unlikely that a model will achieve perfect performance when predicting the expectation component of an individual’s observed outcome. This means that residuals gained from the expectation model will contain prediction error, depending on the model’s accuracy. The ‘error correction’ model allows us to estimate the prediction error for each sample (i.e., the proportion of an achieved residual that is unrelated to CR) and subtract it from the residual (similar to boosting algorithms ^22^). The ‘error correction’ model requires an association between its predictors and the expectation model’s error to predict it. This explains why it did not correct error when we used a linear regression to explain the expectation, as model errors should be normally distributed. By contrast, when using a non-linear model, the error correction improved the achieved CR estimates. Generally, CR estimates made with the inverse learning approach were significantly more accurate and less biased compared to the standard approach across all experiments.

As with any statistical/machine learning approach, sufficient training examples that reflect the expected dynamics must be available to achieve a reliable expectation model. The exact required sample size will depend on the complexity of the data and prediction task. This limitation can be assessed via model validation and managing the bias/variance trade-off^23^. Despite our introduced ‘error correction’ model, achieved CR estimates will likely never be completely error free. The ‘error correction’ model has its own prediction error that will impact the corrected residual, however, as with any predictive model, the reliability of this model can be assessed via validation. Depending on the data and definition what the expectation is, selecting training samples representing this expectation might introduce distribution shifts between the expectation model’s training and application data. Statistical matching could be one approach to mitigate potential distribution shifts^24^.

The inverse learning approach for estimating a CR requires the identification of samples who represent the expected disease dynamics. In the context of estimating CR in AD, this implies that researchers must define the phenotype of ‘non-resilience’. While this is a non-trivial task given the complexity of AD, it also presents a major strength of the inverse learning approach. Employing different definitions of the expectation phenotype provides ample flexibility to investigate specific forms of resilience and requires dedicated thought about the phenotype one aims to describe. For example, individuals following the expectation could be selected based on their clinical progression over time, extracted slopes of cognitive outcomes, cross-sectional cut-offs along several cognitive assessments at different time points, biomarker status and progression, and postmortem pathology. It also allows for splitting based on demographic factors, some of which have been found to influence resilience^25,26^, such as biological sex, race, and ethnicity to derive subpopulation-specific phenotypes of CR. Also, data-driven phenotypes can be leveraged, for example, by using clustering algorithms that partition the data ^2^.

The inverse learning approach will provide more accurate estimates of individual-level resilience under most circumstances. More accurate estimates of CR allow researchers to draw better insights into associations between exposures and CR, their exact effect sizes, and underlying biological mechanisms. These mechanisms are of great interests as they could potentially reveal targets for medical interventions to improve resilience in non-resilient individuals. Also, with respect to personalized medicine, more accurate estimates of an individual’s forecasted resilience could prove beneficial to time the treatment using the newly approved AD drugs^27–29^. Conclusively, understanding resilience and measuring it accurately benefits both, bringing light into the complexity of AD and supporting its management and treatment.

## Data Availability

All data were simulated and the scripts for simulation are available.

https://github.com/Cojabi/inverse_learning

## List of abbreviations

AD: Alzheimer’s disease
CR: cognitive resilience

## Data and code availability

The used data was fully simulated. The code for both the method and data simulation can be found at https://github.com/Cojabi/inverse_learning.

## Acknowledgements Funding

This project is supported by R01AG079142. Coughlan is funded by a NIH Pathway to Independence award, K99AG083063, and an Alzheimer’s Association Research Fellowship, AARF-23-1151259.

