## Supplementary_Material for "Rethinking the residual approach: Leveraging machine learning to operationalize cognitive resilience in Alzheimer’s disease" (from another version of this preprint: version 1, 2024-08-20)

### Simulation non-linear data

Below, we present the non-linear transformation used to simulate a non-linear relationship between predictors, the expectation component, and the cognitive resilience (CR) component of the observed outcome. We used the function as implemented in scikit-learn[1] and presented in Friedman et al.[2]:

$$y(x) = 10 * \sin(\pi * x_1 * x_2) + 20 * (x_2 - 0.5)^2 + 10 * x_3 + 5 * x_4 + noise * N(0,1)$$

### Hyperparameter tuning

We tuned the following hyperparameters in the employed xgboost models using Optuna, a package for Bayesian hyperparameter optimization[3] with the following distributions to sample them from:

- gamma: FloatDistribution(1, 8)
- learning_rate: FloatDistribution(1e-7, 1, log=True)
- max_depth: IntDistribution(3,8)
- subsample: FloatDistribution(0.7, 1, 0.1)

### Schematic comparing CR estimates across residual approaches

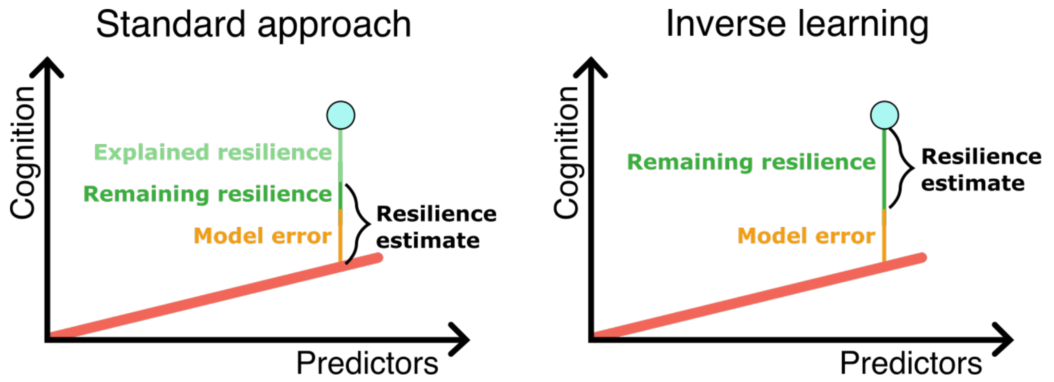


**Figure S1:** Difference between the CR estimates achieved by the standard approach and the proposed inverse learning approach. The standard approach will 'learn away' parts of the resilience component in the observed cognition due to either direct inclusion of associated predictors or correlation. Further, the linear regression will not perfectly predict the observed cognition and thus introduces model error. The residual achieved by the standard approach will thus contain an unknown proportion of error and have lost a proportion of the sought-after information (resilience). The inverse learning approach will not learn any of the predictor-resilience associations and the model error can be quantified and reduced using the 'error correction' model, thus leading to more reliable estimates of cognitive resilience.


1. Pedregosa, F. *et al.* Scikit-learn: Machine Learning in Python. *J. Mach. Learn. Res.* **12**, 2825–2830 (2011).
2. Friedman, J. H. Multivariate Adaptive Regression Splines.
3. Akiba, T., Sano, S., Yanase, T., Ohta, T. & Koyama, M. Optuna: A Next-generation Hyperparameter Optimization Framework. *ArXiv190710902 Cs Stat* (2019).
